# Utility of at-home video recordings for functional skill assessment in Angelman Syndrome: a pilot study

**DOI:** 10.1101/2025.10.06.25337170

**Authors:** Mindy Leffler, Rebecca J. Woods, Amber Sapp, Christina K. Zigler, Robert Komorowski, Rebecca Crean, Lynne M. Bird, Catherine F. Merton, Anna J. Booman, Johnna D. O’Sullivan, Kriszha A. Sheehy, Jessica Duis, Wen-Hann Tan, Anjali Sadhwani

**Affiliations:** Casimir, LLC, Rockville, Maryland, United States of America; Department of Population Health Sciences, Duke University School of Medicine, Durham, NC, United States of America; Ionis Pharmaceuticals, Carlsbad, California, United States of America; University of California San Diego and Rady Children’s Hospital, San Diego, CA, United States of America; Division of Genetics and Genomics, Boston Children’s Hospital, Boston, MA, United States of America; Childen’s Hospital Colorado, University of Colorado, Anschutz Medical Campus, Aurora, Colorado, United States of America; Department of Psychiatry and Behavioral Sciences, Boston Children’s Hospital, Boston, MA, United States of America; Department of Psychiatry, Harvard Medical School, Boston, MA, United States of America; Department of Biostatistics, Harvard T.H. Chan School of Public Health, Boston, MA, United States of America

**Keywords:** neurodevelopmental assessment, video assessment, activities of daily living, real-world assessment

## Abstract

**Background:** Outcome measures currently used to assess function in Angelman Syndrome (AS) may be affected by subject anxiety in clinic, a disconnect between the normed age range of the measure and the study population, and a reliance on caregiver report. This study aimed to develop and pilot a novel outcome for AS, the Angelman Syndrome Video Assessment (ASVA) for the assessment of everyday functional skills in individuals with AS in their home environment.

**Methods:** The task list was informed by published conceptual disease models and determined by a team of experts based on family and clinician input. The study was conducted remotely in the home environment, with families capturing videos of everyday activities and participating in an exit interview about their experience. Videos were evaluated, and caregiver interview transcripts were coded and analyzed to determine whether each task would be included in the finalized measure.

**Results:** Eleven dyads completed the study. The video completion rate was 96%, with 99% of the submitted videos meeting quality standards. Caregivers endorsed the value of assessing people with AS in the home environment. The video capture list was reduced from 27 to 17 tasks.

**Conclusions:** The ASVA is a novel tool that captures data on the daily functioning of individuals with AS in their home environment. This tool presents a potential new way to evaluate subject function in clinical trials and for clinical care.

## Background

Angelman Syndrome (AS) is a neurodevelopmental disorder characterized by severe intellectual disability with minimal or absent speech, epilepsy, and maladaptive behaviors.^1,2^ AS is caused by lack of expression of the maternal copy of *UBE3A* through one of four mechanisms: a deletion of the AS critical region on the maternally-inherited chromosome 15q11.2q13 (∼65%), paternal uniparental disomy (UPD) for chromosome 15q11q13 (∼10%), imprinting defects that result in the maternal allele having the “imprint” of the paternal allele (∼10%), or pathogenic *UBE3A* variants on the maternal allele (∼15%).^1,2^ Those with a deletion are “deletion-positive” while the remaining etiologies are collectively referred to as “deletion-negative”. Those with deletion-negative AS are less developmentally and cognitively impaired than those with a deletion.^,3,4,5,6^ Those with deletion-negative AS have more anxiety and maladaptive behaviors.^7^

In research studies, neurodevelopment of individuals with AS has been assessed using two key clinical outcome assessments (COAs): the Bayley Scales of Infant and Toddler Development (Bayley 3 and 4), an in-person clinician-administered assessment of cognition, language, and motor skills, and the interview version of the Vineland Adaptive Behavior Scales (Vineland 2 and 3), a semi-structured interview conducted by a trained clinician with the caregiver to assess adaptive functioning in the domains of communication, daily living skills, socialization, motor skills, and maladaptive behaviors. ^5,6,7,9,10^ Researchers have also developed the Symptoms of AS-Clinician Global Impression (SAS-CGI) and the Caregiver-reported AS Scale (CASS) as Angelman-specific scales utilizing clinician input and caregiver reports to assess severity of symptoms and changes in symptoms over time.^8^ Since individuals with AS often exhibit high levels of generalized and separation anxiety, especially in unfamiliar environments, their performance on the Bayley in a clinical setting may not be representative of their abilities in daily life. ^9,10,11,12^ In addition, the Bayley is only normed for children up to 42 months, and while it has been used to assess older individuals with significant developmental delays, some of the Bayley tasks may not be developmentally appropriate for adults.^13,14^ The Vineland, the SAS-CGI and the CASS utilize caregiver report of performance, and the accuracy of the data may be impacted by recall bias, the caregiver’s subjective impression of the individual’s ability, and the rater’s interviewing abilities.^15^ ^16,17^ Moreover, the Vineland quantifies how frequently a skill is independently performed and does not capture nuanced qualities of the execution of a skill that may be meaningful to caregivers or the amount of support needed to complete a task.^18^

The inherent limitations in the existing COAs described above call for the need to develop a more objective outcome measure that will allow clinical investigators to assess changes in developmental skills that are meaningful to the caregivers and reflect the true ability of the participants in their natural environments. There is also a need to have an outcome measure that will capture the nuances of the various developmental skills that can only be assessed through direct observations, information not currently captured through the scoring of the Bayley. We therefore sought to develop a performance-based outcome measure in which investigators can use an objective scoring system to rate the level of developmental skills in the different domains by watching video recordings of these individuals performing pre-specified tasks in their natural environments; these ratings would complement the data obtained through the Bayley and the Vineland. By focusing on domains of interest that are important to the families of individuals with AS across all age groups, we aimed to address the unmet need for a relevant and meaningful outcome measure for the entire AS population.

Researchers at Casimir have previously created an outcome measure for DMD to address these limitations, consisting of structured, home-based video captures of everyday movement tasks that are scored by trained and certified central raters utilizing validated scorecards. The Duchenne Video Assessment (DVA) is a performance-based measure that requires a caregiver to record everyday activities in the home environment. The DVA was developed through a structured multi-step process: task selection, video pilot study, scorecard development and validation, and psychometric validation. Details on the development of the DVA have been previously published. ^19,20,21,22^

Given the experience and expertise of the investigators at Casimir obtained through their development of the DVA and the clinical expertise of the investigators at Boston Children’s Hospital (BCH) in Angelman syndrome, the group established a collaboration to develop the Angelman Syndrome Video Assessment (ASVA) in accordance with the guidance provided by the FDA Patient-Focused Drug Development Guidance framework.^23^ Herein, we present the development and piloting of the tasks selected for the ASVA and assess the feasibility of using this measure in the home environment, the acceptability of this measure by caregivers, and the ability of these tasks to capture the different levels of daily functioning skills in the participants.

## Methods

### Participants

Caregiver-participant dyads were recruited for this ASVA Pilot Study, approved by the central IRB IntegReview (Austin, TX), through Boston Children’s Hospital and the Angelman Syndrome Foundation. Eligible participants were required to have a laboratory-confirmed genetic diagnosis of AS and at least 2 years of age to align with the oldest age criteria of clinical trials in Angelman syndrome (ClinicalTrials.gov: NCT05127226 and NCT06914609). Heterogeneous sampling was utilized to include subjects of various ages and molecular subtypes. All caregivers were legal guardians who provided written consent for themselves and the participants.

### Development of the Video Capture Protocol Identifying domains of interest

Extensive prior work had been completed in the AS community to develop patient-centered disease concept models for Angelman Syndrome (AS) to identify what mattered most to caregivers.^24,25^. Two models were developed that were informed by the literature and refined by qualitative interviews with clinicians and caregivers, and they highlighted key domains of interest such as communication abilities, motor function, self-care, executive functioning, seizures, sleep and behavioral challenges. Based on these models, teams from Casimir and BCH focused on capturing functioning in four domains on video: communication, gross motor skills, executive function, and self-care skills as these domains could be evaluated through representative videos. Behavior, sleep, and seizures were excluded due to potential video sampling bias, which could impact symptom quantification.

### Identifying list of tasks

To identify specific tasks to record, the Casimir team conducted semi-structured interviews with six caregivers and two clinicians. Respondents were asked to suggest daily activities within the communication, gross motor, executive function, and self-care domains to assess the functional abilities of individuals living with AS. Additionally, as part of the Angelman Syndrome Natural History Study, the team from BCH had taken initial steps to develop a set of video tasks for the AS population. The team (Table 1) held a series of discussions to evaluate videos of specific tasks previously captured by the BCH team as part of the Angelman Syndrome Natural History Study (ClinicalTrials.gov: NCT04507997), reviewed the list of potential tasks proposed, and determined which tasks should be piloted. Tasks that were foundational to daily life or activities of daily living (ADL) were included if the team determined that video recordings could consistently demonstrate differences in the functional abilities of individuals living with AS. These discussions led to the finalization of 27 tasks across the 4 domains which would be piloted.

**Table 1.**
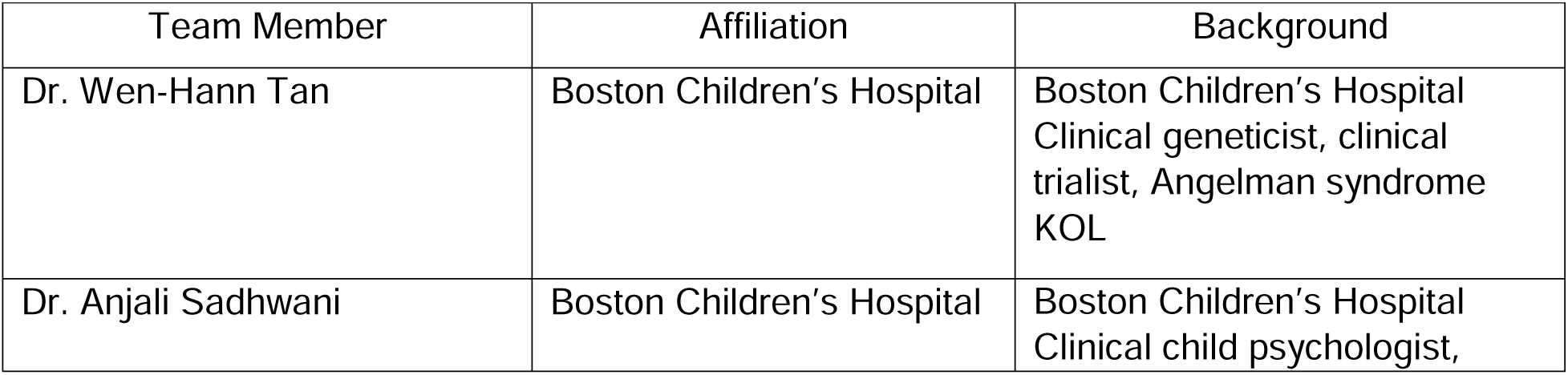
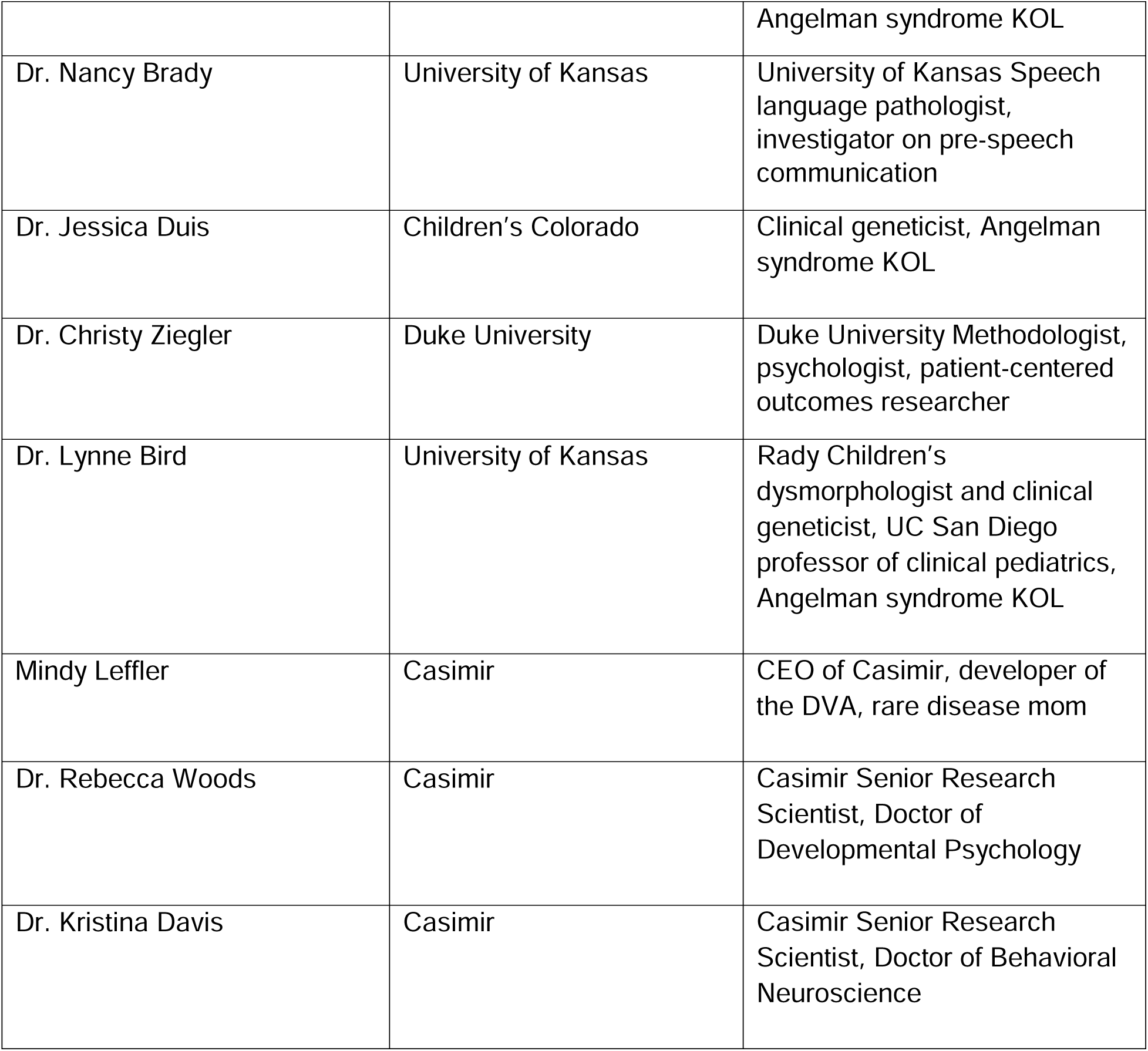
Clinician and Researcher Team (n = 9)

### Developing video capture instructions for the tasks

For each task, video capture instructions were created to help caregivers elicit both mastered and emerging skills. Instructions in the capture procedures specified which aspects of each task needed standardization and which aspects did not, resulting in a draft version of the protocol for the ASVA. Gross motor tasks were categorized based on mobility reported by caregivers regarding each participant’s movements within the home environment. Details of the tasks, task instructions, and materials for each task can be found in the caregiver manual.

### Video Capture

Caregivers recorded participants performing the pre-specified tasks using their personal smartphones or tablet computers using the HIPAA-compliant ShareFile platform (Progress Software Corporation, Burlington, MA). ASVA study kits, including a caregiver training manual, task supplies, a tripod, and a remote-control shutter, were mailed to caregivers. Following receipt, study staff reviewed video capture procedures with caregivers and assisted in downloading the mobile application. Caregivers were instructed to capture an adequate representation of the subject’s abilities and encouraged to record additional videos if initial attempts did not capture the current level of functioning. Caregivers could submit multiple videos per task to include all components. Caregivers were given a two-week window to allow families to record on a day that would allow them to record a representative video of their child’s function. However, families that needed additional time were allowed an additional two weeks because of the large number of tasks and the initiation of the study near the holidays, a busy time for families. Study staff conducted a video quality review within 48 hours of submission to ensure that the overall objective of each activity was accomplished and evaluable behaviors were captured. When necessary, additional recordings were requested to be completed within that additional two weeks.

### Exit Interviews

After submitting the videos, caregivers participated in an exit interview. A semi-structured guide was developed collectively by the Casimir and BCH team members to solicit caregiver’s feedback on their experiences with recording videos, evaluating the clarity of the instructions, and identifying any challenges they experienced. It assessed whether the videos effectively captured the AS participants’ abilities and sought to identify tasks most relevant to family life. Caregivers were also asked for suggestions on improving the ease of recording. Interviews lasted approximately 30 minutes and were conducted over a HIPAA-compliant video conference platform. Interviews were conducted by two PhD research scientists who were part of the Casimir team and were trained on the interview guide and in the conduct of qualitative interviews.

### Evaluation of Video Data

The proportion of tasks completed by each participant was assessed to evaluate the feasibility of each task as well as the overall approach. The appropriateness of the video tasks was evaluated by the Casimir and BCH teams, as well as three additional experts. A speech and language pathologist from Kansas University, a clinical geneticist who serves as the site PI for the Angelman Syndrome Natural History Study, and a psychometrician from Duke University who led the development of the Observer-Reported Communication Ability (ORCA) for Angelman syndrome joined the group for discussion and determination of these tasks. The team reviewed each video using a structured worksheet, noting observations on what was captured, what was observable, and what was scorable for each participant. They also documented deviations in task capture and provided a summary of recommendations for each task.

Overall recommendations from the entire team were compiled for each task. The team deliberated on whether the tasks effectively captured the daily functioning of individuals with AS, whether they highlighted differences in functional abilities between patients, and whether tasks were adequately standardized with consistent capture procedures. Discussions continued until 100% agreement was reached on the inclusion or exclusion of each task.

### Exit Interview Coding

To code the exit interviews, the interview audio was transcribed by a HIPAA-compliant transcription service, and transcripts were de-identified. Transcripts were compared to the audio recording to ensure accuracy. All transcripts were coded by a primary coder, and coding was reviewed by a secondary coder to ensure coding accuracy. Both coders were trained in qualitative research and analysis.

Before reviewing the transcripts, the coders developed a thematic codebook based on the interview guide, and the codes were refined after common themes emerged during the coding process. Coding was performed manually, and text segments were organized by themes to facilitate the calculation of descriptive statistics. Excel was used to track the frequency of codes applied to the transcripts on quantified questions. Key themes were summarized with representative quotes from caregivers and descriptive statistics (n, %).

## Results

### Participants

Eleven caregiver-subject dyads (Table 2) completed the video capture submission and the exit interviews. One caregiver enrolled but did not complete the videos due to a family emergency. Information on the age and molecular subclass of the AS individuals are presented in Table 2. Out of the 11 participants, three were minimally ambulatory and the remaining eight were ambulatory. The participants ranged in age from 4-39 years.

**Table 2.** Demographics of Subjects (n = 11)

| Subject | Sex | Molecular Subclass | Age Range (years) | Ambulatory Status |
| --- | --- | --- | --- | --- |
| 1 | M | UPD | 5-9 | Ambulatory |
| 2 | F | ImpD | Above 20 | Ambulatory |
| 3 | M | Deletion | 5-9 | Ambulatory |
| 4 | M | Deletion | Above 30 | Minimally ambulatory |
| 5 | M | Deletion | 10-14 | Ambulatory |
| 6 | M | Deletion | 15-19 | Minimally ambulatory |
| 7 | M | Deletion | Under 5 | Minimally ambulatory |
| 8 | F | Deletion | 5-9 | Ambulatory |
| 9 | M | UBE3A mutation | 10-14 | Ambulatory |
| 10 | M | Deletion | 5-9 | Ambulatory |
| 11 | F | UBE3A mutation | Under 5 | Ambulatory |

Abbreviations: UPD, uniparental disomy; ImpD, imprinting defect

### Completion of Video Capture

Each dyad was asked to record 20 tasks for each minimally ambulatory subject and 23 tasks for each ambulatory subject (Table 3), resulting in the submission of 339 videos. A quality assurance review of videos revealed that compliance with capture procedures was achieved in 99% (336 out of 339 videos). Three videos were rejected due to failure to follow capture procedures (e.g., not asking a question verbally before providing visual cues). Seven caregivers contacted study staff for assistance or clarification regarding the procedures, with most queries relating to challenges with video upload. The out-of-window submission rate was 26% (89 of 339 videos submitted outside of the two-week window, mean of 9 days past window, 4-15 days range). These videos were included in the determination of the tasks to include in the final measure as the group agreed that the delay in submission would not affect the value of the videos for this purpose.

**Table 3:**
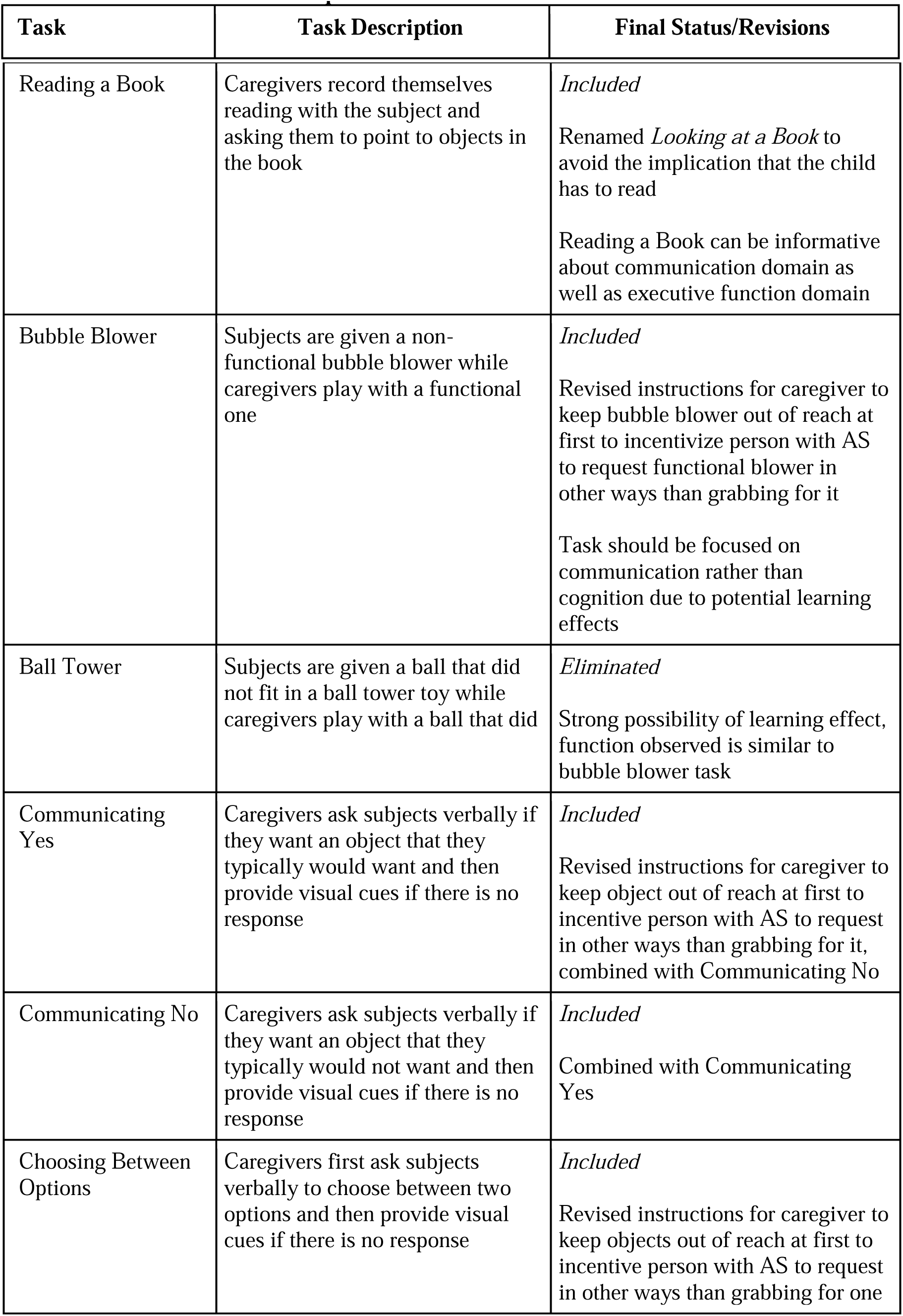
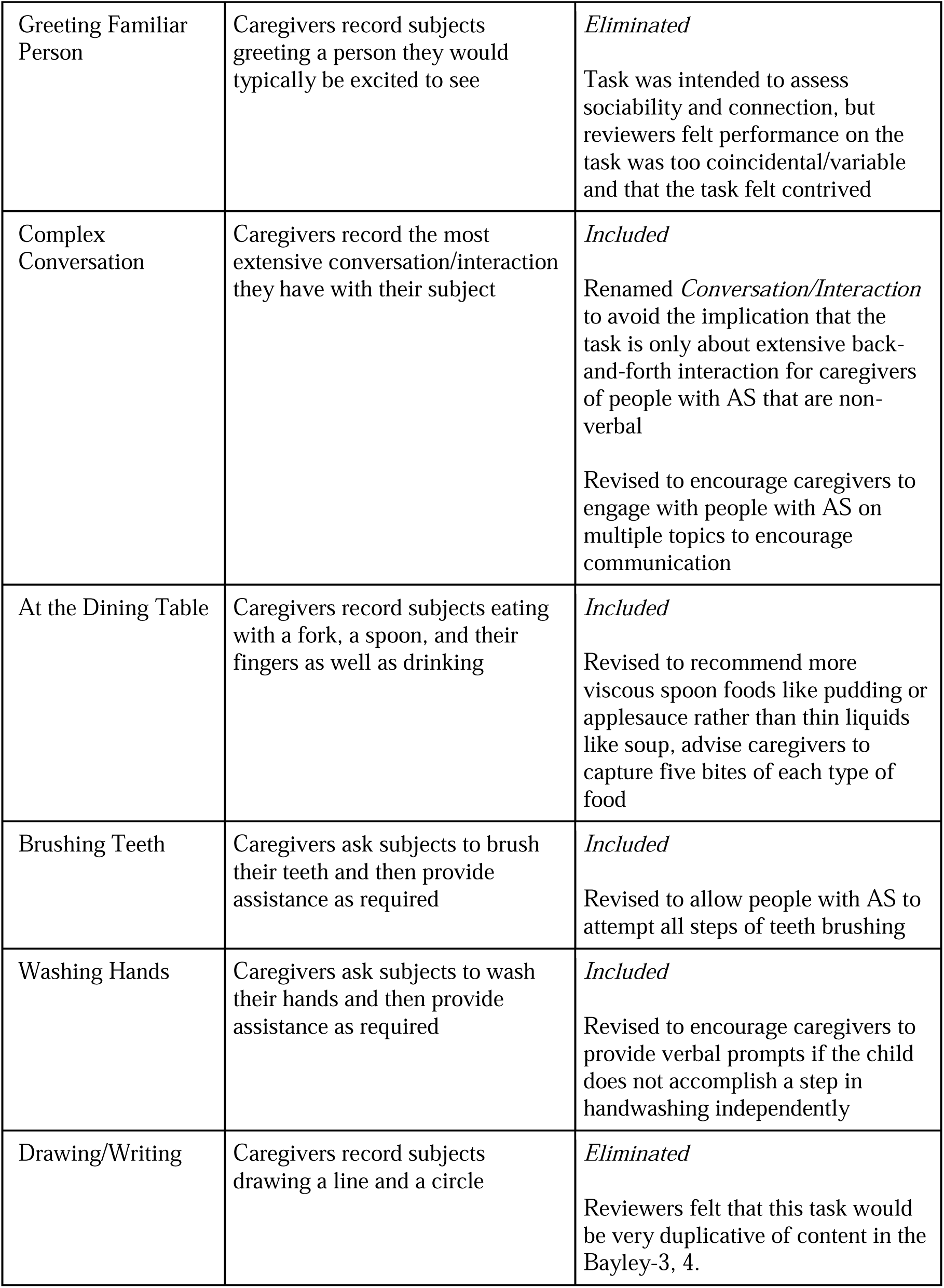
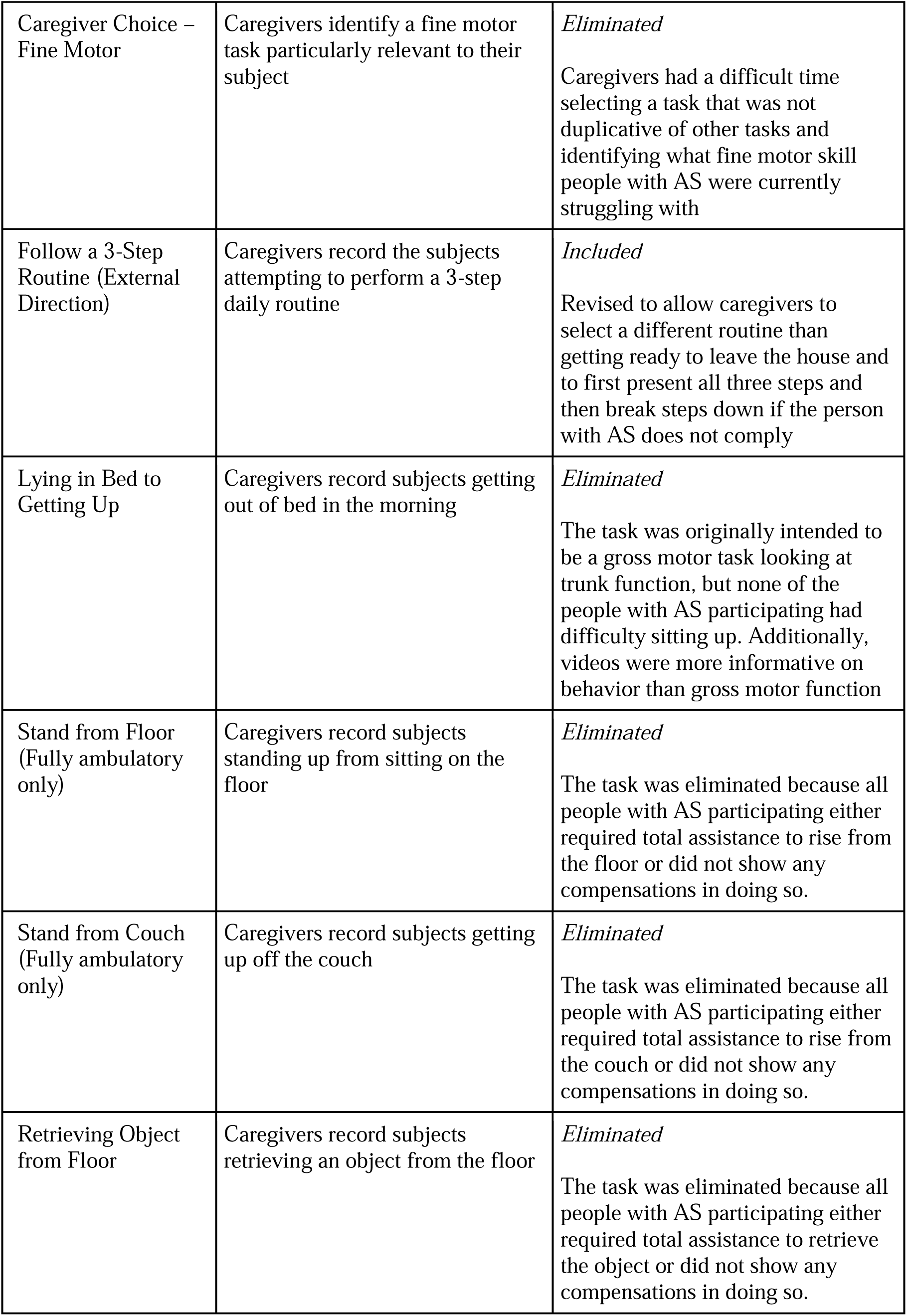
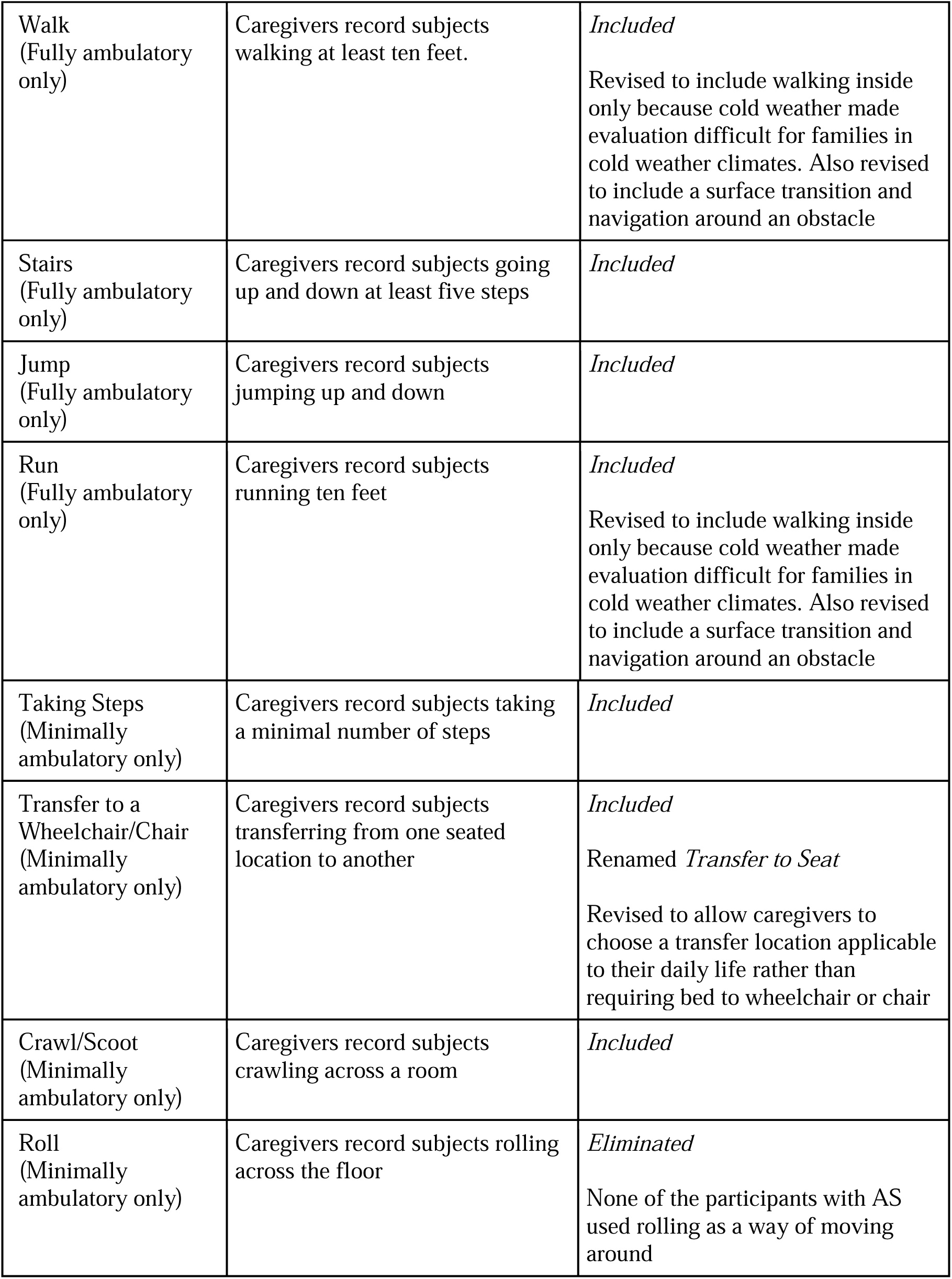
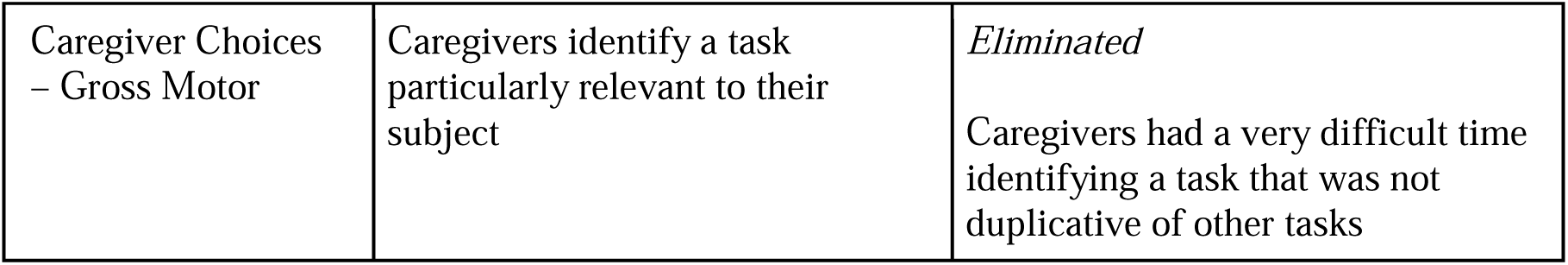
Task List for Video Capture and Final Task Status.

| Task | Task Description | Final Status/Revisions |
| --- | --- | --- |
| Reading a Book | Caregivers record themselves reading with the subject and asking them to point to objects in the book | *Included*<br>Renamed *Looking at a Book* to avoid the implication that the child has to read<br>Reading a Book can be informative about communication domain as well as executive function domain |
| Bubble Blower | Subjects are given a non-functional bubble blower while caregivers play with a functional one | *Included*<br>Revised instructions for caregiver to keep bubble blower out of reach at first to incentivize person with AS to request functional blower in other ways than grabbing for it<br>Task should be focused on communication rather than cognition due to potential learning effects |
| Ball Tower | Subjects are given a ball that did not fit in a ball tower toy while caregivers play with a ball that did | *Eliminated*<br>Strong possibility of learning effect, function observed is similar to bubble blower task |
| Communicating Yes | Caregivers ask subjects verbally if they want an object that they typically would want and then provide visual cues if there is no response | *Included*<br>Revised instructions for caregiver to keep object out of reach at first to incentive person with AS to request in other ways than grabbing for it, combined with Communicating No |
| Communicating No | Caregivers ask subjects verbally if they want an object that they typically would not want and then provide visual cues if there is no response | *Included*<br>Combined with Communicating Yes |
| Choosing Between Options | Caregivers first ask subjects verbally to choose between two options and then provide visual cues if there is no response | *Included*<br>Revised instructions for caregiver to keep objects out of reach at first to incentive person with AS to request in other ways than grabbing for one |
| Greeting Familiar Person | Caregivers record subjects greeting a person they would typically be excited to see | *Eliminated*<br>Task was intended to assess sociability and connection, but reviewers felt performance on the task was too coincidental/variable and that the task felt contrived |
| Complex Conversation | Caregivers record the most extensive conversation/interaction they have with their subject | *Included*<br>Renamed *Conversation/Interaction* to avoid the implication that the task is only about extensive back-and-forth interaction for caregivers of people with AS that are non-verbal<br>Revised to encourage caregivers to engage with people with AS on multiple topics to encourage communication |
| At the Dining Table | Caregivers record subjects eating with a fork, a spoon, and their fingers as well as drinking | *Included*<br>Revised to recommend more viscous spoon foods like pudding or applesauce rather than thin liquids like soup, advise caregivers to capture five bites of each type of food |
| Brushing Teeth | Caregivers ask subjects to brush their teeth and then provide assistance as required | *Included*<br>Revised to allow people with AS to attempt all steps of teeth brushing |
| Washing Hands | Caregivers ask subjects to wash their hands and then provide assistance as required | *Included*<br>Revised to encourage caregivers to provide verbal prompts if the child does not accomplish a step in handwashing independently |
| Drawing/Writing | Caregivers record subjects drawing a line and a circle | *Eliminated*<br>Reviewers felt that this task would be very duplicative of content in the Bayley-3, 4. |
| Caregiver Choice – Fine Motor | Caregivers identify a fine motor task particularly relevant to their subject | *Eliminated*<br>Caregivers had a difficult time selecting a task that was not duplicative of other tasks and identifying what fine motor skill people with AS were currently struggling with |
| Follow a 3-Step Routine (External Direction) | Caregivers record the subjects attempting to perform a 3-step daily routine | *Included*<br>Revised to allow caregivers to select a different routine than getting ready to leave the house and to first present all three steps and then break steps down if the person with AS does not comply |
| Lying in Bed to Getting Up | Caregivers record subjects getting out of bed in the morning | *Eliminated*<br>The task was originally intended to be a gross motor task looking at trunk function, but none of the people with AS participating had difficulty sitting up. Additionally, videos were more informative on behavior than gross motor function |
| Stand from Floor (Fully ambulatory only) | Caregivers record subjects standing up from sitting on the floor | *Eliminated*<br>The task was eliminated because all people with AS participating either required total assistance to rise from the floor or did not show any compensations in doing so. |
| Stand from Couch (Fully ambulatory only) | Caregivers record subjects getting up off the couch | *Eliminated*<br>The task was eliminated because all people with AS participating either required total assistance to rise from the couch or did not show any compensations in doing so. |
| Retrieving Object from Floor | Caregivers record subjects retrieving an object from the floor | *Eliminated*<br>The task was eliminated because all people with AS participating either required total assistance to retrieve the object or did not show any compensations in doing so. |
| Walk<br>(Fully ambulatory only) | Caregivers record subjects walking at least ten feet. | *Included*<br>Revised to include walking inside only because cold weather made evaluation difficult for families in cold weather climates. Also revised to include a surface transition and navigation around an obstacle |
| Stairs<br>(Fully ambulatory only) | Caregivers record subjects going up and down at least five steps | *Included* |
| Jump<br>(Fully ambulatory only) | Caregivers record subjects jumping up and down | *Included* |
| Run<br>(Fully ambulatory only) | Caregivers record subjects running ten feet | *Included*<br>Revised to include walking inside only because cold weather made evaluation difficult for families in cold weather climates. Also revised to include a surface transition and navigation around an obstacle |
| Taking Steps<br>(Minimally ambulatory only) | Caregivers record subjects taking a minimal number of steps | *Included* |
| Transfer to a Wheelchair/Chair<br>(Minimally ambulatory only) | Caregivers record subjects transferring from one seated location to another | *Included*<br>Renamed *Transfer to Seat*<br>Revised to allow caregivers to choose a transfer location applicable to their daily life rather than requiring bed to wheelchair or chair |
| Crawl/Scoot<br>(Minimally ambulatory only) | Caregivers record subjects crawling across a room | *Included* |
| Roll<br>(Minimally ambulatory only) | Caregivers record subjects rolling across the floor | *Eliminated*<br>None of the participants with AS used rolling as a way of moving around |
| Caregiver Choices<br>– Gross Motor | Caregivers identify a task particularly relevant to their subject | *Eliminated*<br>Caregivers had a very difficult time identifying a task that was not duplicative of other tasks |

Caregivers were given the option to omit tasks if the subject was unable to perform a task due to safety or due to other concerns. In total, six dyads chose not to record at least one task: three caregivers with ambulatory subjects did not record running indoors due to limited indoor space, two did not record walking up the stairs due to lack of stairs at home or safety considerations, and one did not record jumping due to safety considerations. Two caregivers with minimally ambulatory subjects omitted rolling and one caregiver omitted crawling because those means of mobility were not routinely used. In total, 96% of the anticipated 244 tasks were completed. Caregivers’ reasons for omitting tasks were documented and considered during the evaluation of individual tasks. This information was used to determine whether to modify the capture procedures, exclude certain tasks from further use, or develop procedures for capturing caregiver rationale for omission that would result in a valid imputation strategy.

### Exit Interviews

Interview transcripts were reviewed for several themes as described below:

#### Attitude towards home based Video Assessment

All caregivers (n=11, 100%) endorsed the need to assess subjects’ daily functioning in their home environment. The primary reason cited was the ability to capture more natural behavior than in the clinic (n=7, 64%). Subject 01 noted: “At the school, whenever they do diagnostic testing,…they think he doesn’t know anything. They think that he can’t do a single thing, because my son won’t do a single thing for him,…the testing environment, he doesn’t know where he is, he’s got a lot of sensory issues, so he can hear every different noise, he knows everything that’s different in the room, why are you making me do all these things, and as far as they’re concerned, my son doesn’t know anything, and I’m like, he knows a whole lot more. He’s actually a lot smarter than you’re able to assess.”

Families also cited the flexibility of the timing of assessment in the home environment (n=4, 36%). Subject 04: “I think it works out much better at home…you can wait for kind of a better time to capture a particular skill.” Caregivers reported that the assessments were fun (n=4, 36%). Subject 12: “A lot of those little activities were really fun for her, and I think they showed her strengths.” Families stated that the familiarity of the home environment provided additional information about their child’s function (n=4, 36%). Subject 02: “In her natural setting, she’ll behave at her…highest level, and not be self-conscious, and… It’s just accurate.” Caregivers also reported that recording videos at home was easier than going to the clinic (n=3, 27%). Subject 03: “We participated in quite a few [studies], and it was probably the easiest one that, as far as like, understanding what we needed to do, and getting it done.”

#### Ability of ASVA to Capture Function

All caregivers (n=11, 100%) reported that the tasks were able to capture their child’s abilities adequately. Subject 01: “These are activities that we would normally do on a regular basis. You know, regular routine, (inaudible) activities, and you know, things like that, you know, they’re just normal play, or normal daily activities.” Some Caregivers reported that their child behaved differently than was typical because of camera distraction (n=2, 18%) or awareness of being watched (n=2, 18%), one caregiver in this group reported that this difference in behavior would result in an underestimation of function (n=10%). Caregivers (n=4, 36%) reported that in-clinic assessment underestimated their child’s abilities for different reasons. Subject 05: “We actually just did the [another AS research study] and they do the Bayley, whatever, Assessment there, and [Redacted - Child’s Name], every time I’m just like, ‘ugh,’ because he is just -- I think he feels penned in this room, he’s getting questions just pounded at him, and it’s… At the end of the day, I don’t think it’s a very accurate assessment of what he’s capable of, because he’s just like, “Do something else, do something else. I want out of here, let’s go.”

#### Clarity of Instructions

All caregivers (n=11, 100%) reported that the procedures and instructions were clear enough to understand. Subject 01: “That instructional sheet? I think it was very helpful, personally I thought it was very helpful having the check off list, was extremely helpful. And then the manual had all the instructions laid out for me. So all, I thought all that was pretty easy. I didn’t have any issues understanding any of that.” Three caregivers reported that the instruction manual was lengthy. Caregivers suggested stating more clearly in the manuals that tasks can be separated into multiple videos, e.g.: one video for feeding self with a fork and one for feeding self with a spoon food (n=2, 18%).

#### Use of Mobile App

Caregivers provided information on the ease of using the mobile application. Caregivers (n=9, 82%) mentioned difficulties getting the videos to upload to the server due to network connectivity issues, phone going into sleep mode during upload, and pace of upload over cellular data connections. One caregiver mentioned the longer videos taking up a large amount of storage space on the phone’s hard drive. Caregivers felt the user interface of the app and upload process made it difficult to know which video covered which activity and which video tasks were complete.

#### Other Difficulties/Suggestions for improvement

Caregivers provided several suggestions to improve the ease of the capture procedures. Caregivers (n=4, 36%) stressed the need for a second person to film or the use of the tripod to allow one person to focus on interacting with the subject. Two families found it difficult to keep siblings out of the video. Caregivers felt some stress over whether they were recording videos correctly (n=4, 36%) or over their child’s awareness of the camera (n=1, 10%). On the video submission timeline, caregivers felt that grouping tasks together into clusters (n=4, 36%) and creating a schedule for families to follow (n=3, 27%) would be helpful. Caregivers also asked for clarification that recording times are suggestions rather than requirements (n=2, 18%) and guidance on what to do if a child cannot perform the task (n=2, 18%). Subject 03: “I was going through each video, they -- some of them were like, record for like, five minutes. And I’m like, my son doesn’t do anything for more than 30 seconds.”

#### Most/Least Relevant Tasks

Caregivers provided input on the tasks they felt highlighted their child’s abilities in a manner that was relevant to daily life and identified tasks that they felt did not. Rather than commenting on each task in turn, caregivers interpreted this question as a request to mention the tasks that were the most or least relevant to their family. The *At the Dining Table* and *Communication* tasks were noted to be particularly relevant by caregivers (n=6, 55%). Subject 03: “I think any of the communication videos were…important, especially for our family.” Subject 05: “So I felt like just all the ones about communicating were more difficult, but at the same time I feel like we got a really good like the videos that we did get, I think, were pretty good at conveying how he communicates, you know, and what it’s like.” Each task was endorsed by a similar number of caregivers (1-3) as being the most “relevant”. A similar trend was observed in the tasks that caregivers considered to be not particularly relevant to the subject’s current level of functioning, with the *ball tower* (n=4, 36%) and the *crawling task* (n=3, 27%) being mentioned by the most caregivers. No other task was rated as irrelevant by more than two caregivers.

### Revision of Video Assessment

Based on the team’s evaluation of the video data and results from exit interviews, several changes were made to the final task list. Eleven tasks were eliminated from the assessment –five tasks from the communication, executive functioning, and ADL domains and eight in the gross motor domain. Within the gross motor domain, tasks such as rolling (which none of the AS indivduals could do) and standing up from the couch (which everyone needed support for) were removed. Thirteen tasks were revised. Revisions included simplifying instructions, focusing the task on areas of difficulty for individuals living with AS, and increased standardization. Suggestions from exit interviews and from common procedural errors identified in videos were used to edit capture procedures for clarity. The final version of the ASVA was revised andincluded 14 tasks for those who were fully ambulatory and 11 for those who were minimally ambulatory. The reasons for elimination and revision are detailed in Table 3.

Revisions were also made to the capture procedures and caregiver training based on the results of the exit interviews. Caregiver training materials were revised to suggest the need for a second adult to record or the use of a tripod. Training materials were also revised to emphasize the intent of the videos to capture subject’s current level of function accurately, rather than providing a “test for them to pass”, so caregivers should not feel concerned about submitting videos that documented inability to perform a task. Caregiver training videos were produced to complement the training manuals for caregivers who requested a demonstration of procedures. Tasks were grouped for more convenient recording, and a schedule was added to guide caregiver recording efforts.

## Discussion

This study developed and piloted a home-based video assessment (ASVA) for individuals with AS across a wide age group with varying developmental abilities. The study team developed a series of tasks to assess functioning in four core domains, viz. communication, gross motor skills, executive functioning, and self-care skills through video recordings of prespecified tasks in the home environment with standardized instructions. Eleven caregiver-patient dyads piloted the measure. Overall, there was strong support for the feasibility and acceptability of the measures, with high compliance rate. They strongly endorsed the use of home video recordings to better understand the functioning of their child in a naturalistic setting. In addition, caregivers appreciated the clarity of the instructions and confirmed the ability of the ASVA tasks to adequately capture participants’ functioning. Based on caregiver feedback and expert review of piloted videos, the final ASVA task list was revised, resulting in the elimination of certain tasks and modification of instructions for the tasks and the video capture procedures.

A survey of the literature revealed a need for AS-specific outcomes that accurately capture daily functioning and do not rely on caregiver recall in observational studies and clinical trials.^26,27^ The ASVA has the potential to meet this unmet need by and providing a structured mechanism to assess individuals with AS in a familiar environment, gaining insight into their functioning in daily life. Remote assessment can reduce the burden of study participation for families that already face high care burden^28,29^ It also has the potential to enhance accessibility to clinical studies, thereby improving equity and inclusion in these studies. Furthermore, the ability to blind central raters of the videos to the timepoint in clinical trials has the potential to reduce rater bias. Video capture also allows raters to review the recording repeatedly, which potentially allows for greater accuracy of ratings.

Feedback provided by caregivers and review of videos submitted enabled the study team to reflect on the potential limitations of the ASVA and take mitigation steps to revise the tasks and instructions. To address the concern of caregiver-subject burden, tasks was reduced, shortened, or simplified. Many caregivers did not find the ShareFile mobile application intuitive; future studies will utilize a custom application designed for ease of video capture and submission that integrates training alongside capture. Furthermore, caregivers were made aware of the possibility of extending study windows at the start of the study. Setting firmer deadlines might have reduced window extensions. Lastly, distraction of the participant by the smartphone or tablet was observed, which may be mitigated with discrete placement of the tripod and use of a remote shutter or recording by a second adult.

We acknowledge several limitations of this pilot study. Initial selection of domains was determined based on prior work on the conceptual disease model in the AS community; however, only a small number of clinicians and caregivers were interviewed to brainstorm the initial video capture task list within the domains identified by the conceptual model. Only 11 AS families completed the study, and only two AS individuals were over 18 years of age, limiting our ability to determine whether this would be useful and acceptable to the caregivers of these adults. The exit interview guide question that asked caregivers to identify tasks highlighting their child’s abilities as being relevant or irrelevant to daily life should have prompted caregivers to assess the relevance of each task individually rather than relying on them to recall tasks spontaneously from memory. A potential criticism of the development of the ASVA is the lack of an *a priori* threshold for the inclusion or exclusion of each task in the final task list. However, setting thresholds for task elimination, such as a percentage of caregivers that identified the task as difficult or a percentage that omitted the task, may have been too restrictive to guide decision-making for such a heterogenous population and would not allow investigators to account for capture procedure revisions that could help individual tasks meet thresholds. Therefore, we relied on the clinical judgment of experts in the AS community to reach consensus on which tasks would be eliminated or retained.

The next steps in the development of the include the launch of a larger study in individuals with AS and neurotypical controls that will generate video data on the finalized ASVA tasks. These videos will be reviewed and used for the development and validation of scorecards that quantify function. A Delphi panel will be convened to obtain consensus on the meaningfulness, clarity, and comprehensiveness of the scoring methodology, and the psychometric properties of the ASVA will be evaluated.

## Conclusions

The ASVA is a new outcome measure designed to allow for the evaluation of daily functioning in individuals with AS in their home environment This study found that the approach is feasible in families with AS. Expert reviewers finalized a video task list that targeted domains validated by conceptual models in AS and resulting videos provided evaluable information on the function in individuals with AS. Future work includes expanding the range of videos to be collected and developing and validating scorecards for standardized rater assessment of video capture data.

## Data Availability

All data produced in the present study are available upon reasonable request to the authors

## Declarations

### Ethics approval and consent to participate

This study received ethical approval from IntegReview, a central Institutional Review Board, and the study was performed in accordance with the ethical standards as laid out in the 1964 Declaration of Helsinki and its later amendments. Approval number CAS-CAS004-01, approved December 31, 2020. Clinical trial number: not applicable.

### Consent for publication

Not applicable.

### Availability of data and materials

The data that support the findings of this study are not publicly available due to privacy or ethical restrictions. The data are available on request from the corresponding author.

### Authors’ contributions

Authors M.L., R.W., A.S., C.Z., R.K., R.C., L.B., J.D., W.T., and A.S., conceived, designed, conducted, analyzed, and interpreted the results of the study. C.M., A.B., and J.O., collected data. K.S. assisted in interpretation of the data. Authors M.L. and R.W led manuscript writing. All authors discussed results, reviewed the manuscript for important intellectual content, provided feedback, contributed to, and approved the final manuscript.

## Acknowledgements

The authors are grateful to the patients and caregivers who participated in this study.

